# Supplementation with short-chain fatty acids and the prebiotic 2FL improves clinical outcome in PD

**DOI:** 10.1101/2023.11.01.23297866

**Authors:** Tobias Hegelmaier, Alexander Duscha, Christiane Desel, Sabrina Fuchs, Michal Shapira, Qihao Shan, Gabriele I Stangl, Frank Hirche, Stefan Kempa, András Maifeld, Lisa-Marie Würtele, Jana Peplinski, Diana Jauk, Claudia A. Dumitru, Ute Obermüller-Jevic, Svein-Olaf Hustvedt, Nina Timmesfeld, Ralf Gold, Antonia Zapf, Ibrahim E. Sandalcioglu, Sanaz Mostaghim, Horst Przuntek, Eran Segal, Nissan Yissachar, Aiden Haghikia

**Affiliations:** Department of Neurology, Otto-von-Guericke University, Magdeburg, Germany; German Center for Neurodegenerative Diseases (DZNE), Magdeburg, Germany; Department of Molecular Cell Biology, Weizmann Institute of Science, Rehovot, Israel; Department of Computer Science and Applied Mathematics, Weizmann Institute of Science, Rehovot, Israel; Faculty of Computer Science, Otto-von-Guericke University Magdeburg, Magdeburg, Germany; Institute of Agricultural and Nutritional Sciences, Martin-Luther-University Halle-Wittenberg, Halle (Saale), Germany; Integrative Metabolomics and Proteomics, Berlin Institute of Medical Systems Biology/Max Delbrück Center for Molecular Medicine, Berlin, Germany; Department of Neurology, Ruhr University Bochum, St. Josef Hospital Bochum, Bochum, Germany; Department for Neurosurgery, Otto-von-Guericke University, Magdeburg, Germany; Nutrition & Health Division, GBU Nutrition Ingredients, BASF SE, Ludwigshafen, Germany; Nutrition & Health Division, GBU Pharma Solutions, BASF A/S, Oslo, Norway; Department for Medical Informatics, Biometry and Epidemiology, Ruhr-University, Bochum, Germany; Institute of Medical Biometry and Epidemiology, University Medical Center Hamburg-Eppendorf, Hamburg, Germany; Clinic of Neurology II, EVK Hattingen, Hattingen, Germany; The Mina and Everard Goodman Faculty of Life Sciences, Bar-Ilan University, Ramat-Gan, Israel; Bar-Ilan Institute of Nanotechnology and Advanced Materials, Bar-Ilan University, Ramat-Gan, Israel

## Abstract

**Background:** Parkinson’s disease (PD) is associated with dysbiosis, proinflammatory gut microbiome, disruptions to intestinal barrier functions, and immunological imbalance. Microbiota-produced short-chain fatty acids promote gut barrier integrity and immune regulation, but their impact on PD pathology remains mostly unknown.

**Objectives:** To evaluate supplementation with short-chain fatty acids as an add-on intervention in PD.

**Methods:** In a randomized double-blind prospective study, 72 PD patients received short-chain fatty acids and/or the prebiotic fiber 2′-fucosyllactose supplementation over 6 months.

**Results:** We observed improvement in motor and nonmotor symptoms, in addition to modulation of peripheral immunity and improved mitochondrial respiration in immunocytes. The supplementation had no effect on microbiome diversity or composition. Finally, multiobjective analysis and comprehensive immunophenotyping revealed parameters associated with an optimal response to short-chain fatty acids and/or 2′-fucosyllactose supplementation.

**Conclusion:** Short-chain fatty acids ameliorate clinical symptoms in Parkinson’s disease patients and modulate mitochondrial function and peripheral immunity.

## INTRODUCTION

Parkinson’s disease (PD) is one of the most common progressive systemic neurodegenerative disorders, affecting millions of people worldwide. Despite intensive research, the cause of neurodegeneration is not fully understood. The current state of research assumes a multifactorial etiology. In addition to sporadic forms of genetic predisposition, mitochondrial dysfunction, oxidative stress and neuroinflammation, also environmental factors play a crucial role.^1^ Gastrointestinal symptoms are often the first nonmotor symptoms in PD, in addition to olfactory dysfunction, which in most cases occur years to decades before the first motor symptoms, i.e., rigor, tremor and akinesis.^2^ PD incidence rates are rising in addition to changes in nutrition and the consumption of Western-style diets, which are low in fiber and high in saturated fats and refined carbohydrates. These dietary habits lead to a dysbiotic intestinal microbiota and altered metabolome as well as intestinal inflammation.^3^ Growing evidence supports the idea that microbial dysbiosis, leaky gut syndrome and a proinflammatory intestinal environment are central components of the pathogenesis of PD.^4–7^

The gut metabolome is essential for metabolic homeostasis but is also involved in communication between the microbiome and the subepithelial structures of the gut, which has multifold systemic implications for the organism. Short-chain fatty acids (SCFAs) are a major group of metabolites involved in the microbiome-gut interaction and are produced through the anaerobic fermentation of dietary fibers. A large proportion of SCFAs are metabolized by colonocytes or in the liver, where they contribute to the energy supply. However, a broad range of intracellular SCFA effects are also mediated by G-protein-coupled receptors and SCFA transporters. These include maintenance of intestinal barrier integrity, mucus production, protection against intestinal and systemic inflammation and blood‒brain barrier (BBB) permeability.^8^ In PD patients, the levels of SCFA-producing bacteria and fecal SCFAs are significantly reduced.^9^ In a recent study investigating the potential therapeutic effect of propionate (PA), we observed a putative neuroprotective effect in addition to immune regulation.^10^ Targeting the gut-brain axis via the microbiome, metabolome and intestinal barrier is a promising approach for future therapies for PD. Consumption of prebiotic fibers has recently been tested in a small cohort of PD patients over 10 days.^11^ However, SCFA supplementation in PD patients over a prolonged period has not yet been evaluated.

Here, we conducted a randomized, double-blind prospective study over 6 months to investigate direct supplementation of the SCFAs PA and butyric acid (BA) and the prebiotic 2′-fucosyllactose (2FL).^12^ The primary endpoints of this study were the impact on microbiome diversity and composition as well as changes of SCFA concentration in stool and serum. Because the study was exploratory, no correction for multiplicity and no confirmatory statements are made. The secondary endpoints were the effect on the clinical parameters defined by The Movement Disorder Society-Sponsored Revision of the Unified Parkinson’s Disease Rating Scale (MDS-UPDRS III), levodopa equivalent daily dose (LEDD), PANDA, and olfactory score. We identified novel mechanistic connections between SCFA supplementation, systemic immunity and clinical outcomes in PD patients.

## METHODS

### Study design and analysis

The study was performed from November 2019 to August 2020 after being approved by the Ethics Committee of the Ruhr-University Bochum (November 2019; registration number 19-6713). Prior to participation, all subjects signed informed consent forms. This clinical trial was registered in the German Clinical Trials Register (DRKS; registration number DRKS00027061). A total of 94 participants were assessed for eligibility and 72 were confirmed for randomization and assigned to one treatment group upon recruitment. Randomization was performed by permuted block randomization to one of the three study arms. The randomization list was prepared by BASF SE and provided to the hospital pharmacy in a sealed envelope. All patients were instructed to take either 3810 mg PA+BA capsules (BA: 2550 mg; PA: 1260 mg) with 2400 mg placebo, 3250 mg 2FL capsules with 3200 mg placebo or 3250 mg 2FL capsules with 3810 mg PA+BA capsules daily for up to 6 months in combination with existing PD-specific therapy. The PA+BA, 2FL and placebo were provided as delayed-release capsules by BASF SE. Individual subjects were seen at the outpatient clinic every 3 months after initiation of supplementation and underwent complete neurological assessment performed by a certified Neurostatus C neurologist. Blood and fecal samples were collected at the Department of Neurology, St. Josef Hospital Bochum, at the Clinic of Neurology II, EVK Hattingen, and at the University Clinic of Neurology, UK Magdeburg. Clinical data were evaluated by 2-way ANOVA with time and treatment as factors or mixed effects analysis with patient as random effect. It was not corrected for multiplicity, so the p-values are used as descriptive measures. Fecal samples were immediately frozen at -80°C. Serum tubes were centrifuged at 30 min after the blood draw, and the supernatant frozen at -80°C. For isolation of PBMCs, blood was drawn in EDTA tubes and separated using Cytiva Ficoll-Paque™ PLUS. Isolated cells were frozen in CTL-Cryo medium (Immunospot) and stored at -80°C.

### Quantification of SCFA levels in serum and fecal samples

Methods of sample preparation were previously described.^10^ SCFA levels were measured by HPLC‒MS/MS (Agilent 1100 HPLC).

### Metagenomic sequencing and bioinformatics analysis

Metagenomic DNA was extracted using the DNeasy PowerMag Soil DNA extraction kit (Qiagen). Next-generation sequencing (NGS) libraries were prepared using Illumina’s Nextera DNA library prep and sequenced on an Illumina NovaSeq sequencing platform with 100 bp single-end reads at a depth of 10 million reads per sample. Reads containing Illumina adapters and low-quality reads were filtered out, and ends of low-quality reads trimmed. Reads were mapped to the human genome using bowtie with inclusive parameters, and matches were discarded. The relative abundance of bacterial species was obtained by an expanded microbial genome reference,^13^ with default parameters. Microbiome α-diversity was calculated by the Shannon diversity index. Richness was calculated as the number of species in the sample detected with an abundance of at least 1e-4. All abundances were logarithmically transformed. To evaluate the discriminative power of microbial composition for response prediction, we developed an XGBoost^14^ prediction model that exclusively utilizes microbiome features as inputs and outperforms other methods for human microbiome data classification.^15^ The receiver operating characteristic (ROC) curves mean and standard deviation were calculated using the curves produced in a fivefold cross-validation approach. To ensure the model’s robustness, we conducted a label-swapping analysis to determine that the performance of the model was no better than random prediction, resulting in an area under the curve (AUC) value very close to 0.5. To gain insights into the model’s interpretability, we utilized SHAP (SHapley Additive explanation)^16^ to analyze feature attributes. The SHAP values represent the average change in the model’s output when conditioning on a specific feature.

### Immunophenotyping

For in-depth immunophenotyping archived PBMCs were stained according to standard protocols. Antibodies used and gating strategies applied are listed in Supplementary Tables S1,S2. Staining panels and gatings were modified based on Monaco et al.^17^ Data was recorded on a BD FACSCelesta ^TM^ with standardized application settings.

### Treg *in vitro* assays

Treg suppression assays were performed as previously described^10^ using whole blood from a validation cohort of PD patients at baseline and after 14 days of supplementation with 2FL or BA+PA. For Treg/PBMC cocultures with *in vitro* addition of BA and PA, blood from healthy controls or PD patients without SCFA supplementation was used. Treg/PBMC cells were isolated as described and seeded on a 1:1 ratio in serum free TheraPEAK^TM^ X-VIVO^TM^ 15 (Lonza) without CFSE staining. Cells were stimulated with 5 μg/ml PHA (Phytohemagglutinin, Merck), 150 μM BA (Merck) and 150 μM PA (Merck) for 4 days. Cell culture supernatants were analyzed using a LEGENDplex^TM^ Human Essential Immune Response Panel (BioLegend) and recoded on a BD FACSCelesta^TM^.

### qPCR analysis of sorted Tregs

PBMCs from the validation cohort were isolated and 5x10^7^ cells stained with αCD4-FITC (RPAT4, BD Biosciences), αCD25-APC (BC69, BioLegend) and αCD127-PE (HIL-7R-M21, BD Biosciences). CD4^+^CD25^++^CD127^lo^ Tregs were sorted on a BD FACSAria^TM^ III. RNA was isolated using an RNeasy Micro kit (Qiagen) and transcribed using a QuantiTect® Reverse Transcription Kit (Qiagen). QPCR was performed on a QuantStudio^TM^ 7 Real-Time PCR system using TaqMan® Fast Advanced Master Mix and TaqMan® Gene Expression assays (Applied Biosystems): *B2m*: Hs00187842_m1, *Ccr8*: Hs04969449_m1, *Cmc1*: Hs00976539_g1, *Crot*: Hs00221733_m1, *Ctla4*: Hs00175480_m1, *Foxp3*: Hs01085834_m1, *Il10*: Hs00961622_m1, *Il17rb*: Hs00218889_m1, *Xpa*: Hs00902270_m1.

### Seahorse XF Cell Mito Stress Test

Cellular metabolic activity was measured using a Seahorse XFp Cell Mito Stress Test Kit (Agilent Technologies) on a Seahorse XFp analyzer according to the manufacturer’s protocol. Frozen PBMCs were thawed and 3x10^5^ cells per well were seeded on poly-D-lysine-coated 6-well Agilent Seahorse XF Cell Culture Microplates in X-VIVO^TM^ 15 (Lonza). Cells were incubated for 24 h at 37°C prior to Seahorse assay performance. Individual parameters were calculated according to the Seahorse XFp Cell Mito stress test protocol.

### Multiobjective analysis

Multiobjective analysis (MOA) is based on the concept of multiobjective optimization and is applied to problems with several conflicting objectives, where increasing the quality in one objective results in deteriorating the others. Multiobjective problems usually contain a subset of data points that are not dominated by others in the dataset and are located on a front surface if mapped to the objective space. The central idea concerns the nondominated sorting of a dataset^18^ and then clustering the data according to the so-called fronts. The clinical dataset *A* of N patients, was sorted according to the domination criterion as follows: A data point X dominates a data point Y given the objectives f1 = PANDA, f2 =MDS-UPDRSIII and f3 = olfactory score if:

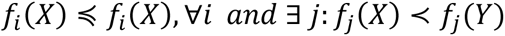

where < and ≤ denote better and equal or better, respectively.

By applying the domination criterion to the dataset *A*, a subset of data points that are not dominated by any other was identified. This subset was indicated as Front 1. Next, the same procedure was performed on dataset *A* without the data points in F1. By performing this procedure iteratively, the dataset was sorted into several fronts from F1 (best subset) to Fw (worst subset). For further analysis we took 20% of patients with the lowest and highest front numbers and sorted them into clusters *A*_1_ and *A*_2_. For binary correlation-based feature selection the ratio ln(V2/baseline) for every determined physiological parameter was calculated and converted entries into rankings. Rankings were normalized (mean of 0 and standard deviation of 1) and the mean and standard deviation of the parameters was computed within each cluster. In this case, the mean describes the level of deviation of a particular parameter within a cluster from the rest of the population, while the standard deviation describes the level of concentration compared to the rest of the population.

### Data Sharing

This study did not generate new unique reagents. All new datasets will be made freely available at the time of publication. Any additional information required to reanalyze the data reported in this work is available from the corresponding author upon request. This paper does not report any original code or algorithms. The code used in this study is freely available at https://www.ci.ovgu.de/Research/Codes.html and on GitHub.

## RESULTS

### Improved clinical outcome upon 6 months supplementation in PD patients

A total of 72 patients were included and randomly assigned to receive 2FL+placebo, PA+BA+placebo or a combination of 2FL+PA+BA. Study visits with detailed neurological examination and sample collection were performed at the beginning of the study (baseline) and after 3 (Visit1, V1) and 6 months (Visit2, V2) (Supplementary Fig. S1). Study groups were comparable in terms of baseline variables (Supplementary Table S3). Supplementation was generally well tolerated. One patient in the 2FL group discontinued treatment due to a mild nontreatment-associated adverse event, hence no follow up data are available.

We assessed extrapyramidal motor function using the MDS-UPDRS III^19^ and LEDD^20^, olfactory function using the Sniffin’ Sticks test^21^ and cognitive function using the PANDA.^22^ All 3 interventions showed a decrease in MDS-UPDRS III scores and LEED over the period of 6 months (Fig. 1A, B). Improvement in olfaction, however, was observed only in the 2FL and combination group 2FL+BA+PA (Fig. 1C), while PANDA revealed an improvement in cognitive functions in all groups (Fig. 1D). In sum, 6 months of supplementation with SCFA and/or 2FL improved motor function, led to a reduction in the LEDD required and positively impacted olfactory and cognitive function in patients with PD.

**Fig. 1.**
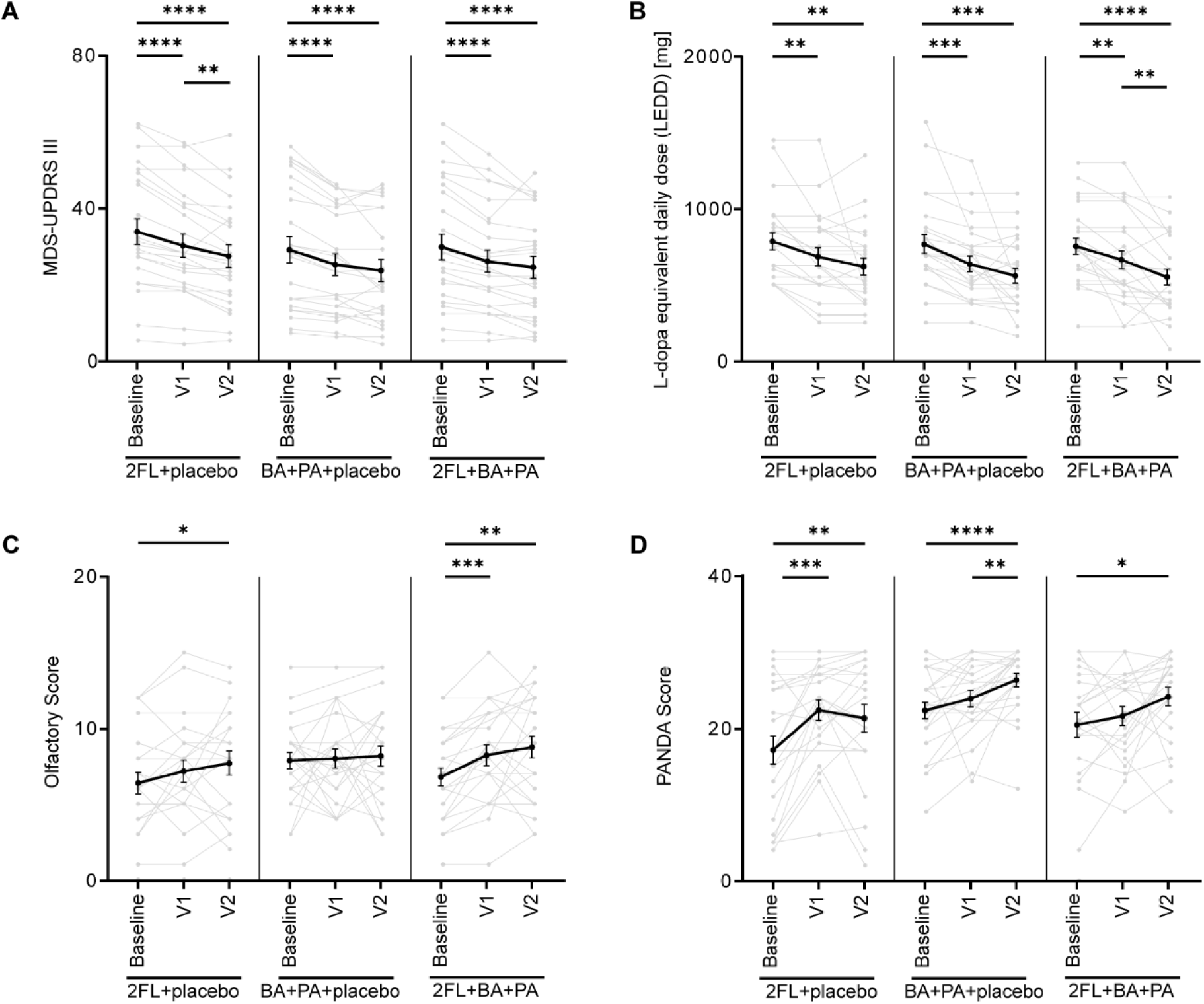
Improved clinical outcome upon supplementation. Clinical parameters determined at the first study visit (Baseline) and at the three (V1) and six (V2) month follow-ups. (**A**) MDS-UPDRS III, (**B**) LEDD, (**C**) olfactory score and (**D**) PANDA. Data plotted as before-after for each patient (gray) and mean (black) ± SEM. Tested by 2-way ANOVA (C, D) or mixed effects analysis (E, F). **** p<0.0001, *** p< 0.001, ** p < 0.01, *p < 0.05.

### Gut microbiome diversity is not altered following supplementation

We collected stool samples from all participants and performed shotgun metagenomic sequencing. Supplementation did not change microbiome diversity or richness in any of the study groups (Fig. 2A,B) and concentrations of SCFA in fecal or serum samples were not elevated (Supplementary Fig. S2). Thus, consistent with our previous study^10^, SCFA and/or 2FL supplementation did not result in measurable alterations to gut microbiome composition.

**Fig. 2.**
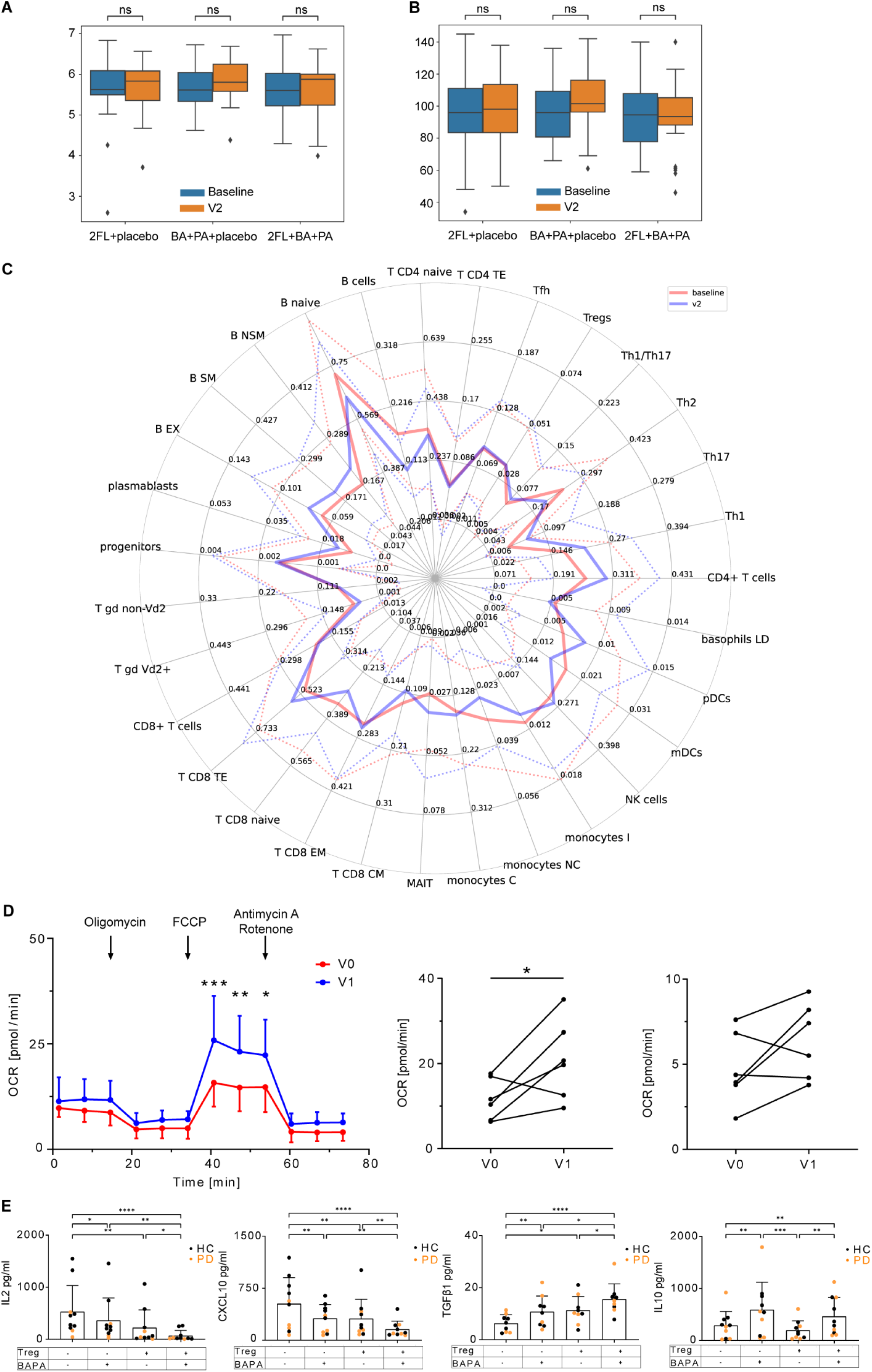
Effect of supplementation on the gut microbiome and immune cells. Shotgun metagenomic sequencing of n=71 stool samples. (**A**) Shannon diversity, (**B**) richness of gut microbiota at baseline and V2. (**C**) Ratio of immune subsets at baseline (red) and 6 months (V2, blue). Mean (solid line) ± SD (dashed lines), all interventions pooled. (**D**) Mitochondrial respiration; validation cohort at baseline (V0) and after 14 days of (V1, n=3 2FL or BA+PA). Left: Oxygen-consumption rate (OCR) over time, middle: Maximal and right: Basal respiration, mean ± SD, significance determined by T test. (**E**) Treg/PBMC coculture assay of n=5 healthy controls (HC) or PD patients, *in vitro* addition of BA+PA. Data presented as mean ± SD, significance determined by Friedman multiple comparisons test.

### SCFA/2FL supplementation modulates immune cell subsets

SCFAs have been shown to exert immunomodulatory effects;^10, 23^ therefore, we performed in-depth characterization from cryopreserved peripheral blood mononuclear cells (PBMCs). We detected changes in cell proportions across almost all subsets analyzed (Fig. 2C). Thus, supplementation with 2FL and/or SCFA induces multifaceted alterations in the composition of the peripheral immune compartment.

Since we have previously shown that SCFA improves mitochondrial respiration and restores the suppressive function of regulatory T cells (Treg) in patients with multiple sclerosis (MS)^10^, we recruited a validation cohort with n=4 PD patients receiving either 2FL or BA+PA. Maximal respiration was significantly elevated after 14 days of supplementation (Fig. 2D), suggesting that the intervention positively affected mitochondrial function. We did not detect any changes in Treg suppressive capacity, but sorted Tregs showed an increase in the expression of the mitochondrial genes (Supplementary Fig. S3A-C). We further assessed the functionality of Tregs by coculture experiments with and without the addition of BA+PA *in vitro*. Secretion of pro-inflammatory CXCL10 and IL2 was significantly decreased, while the levels of anti-inflammatory TGFβ and IL10 increased (Fig. 2E). In summary, SCFA supplementation improves mitochondrial respiration in immunocytes, inhibits the release of proinflammatory mediators and increases the secretion of anti-inflammatory mediators from immune cells.

### Distinct changes in immune cell subsets in responding patients

While 6 months of supplementation improved motor function, this was not observed in all patients. In every intervention group, there were few patients with unchanged or increasing MDS-UPDRS III scores over time (Supplementary Fig. S3D). To gain insight into the mechanisms underlying successful SCFA intervention, we combined all 3 intervention groups and stratified them into responders (R: MDS-UPDRS III V2 < MDS-UPDRS III baseline) and nonresponders (NR: MDS-UPDRS III V2 ≥ MDS-UPDRS III baseline). We detected significant increases of CD4 T cells after 6 months of intervention (Fig. 3A). Supplementation differentially affected subsets of CD4 T cells, with significantly increased Th1- and Th17 cells in the R and NR groups, while the levels of Th2 cells were significantly reduced only in responding patients. The levels of B cells were also significantly reduced both in the R and NR groups (Fig. 3B). Remarkably, the percentages of mucosal-associated invariant T cells (MAIT), nonclassic and intermediate monocyte subsets only significantly changed in responders (Fig. 3B). The percentages of other immune cell subsets were not significantly altered. In summary, supplementation with 2FL and/or SCFA induces a multitude of changes in the composition of immune cell subsets in the blood regardless of treatment response, while increased numbers of MAIT cells and decreased numbers of nonclassic and intermediate monocytes seem to correlate exclusively with intervention-induced clinical improvement.

**Fig. 3.**
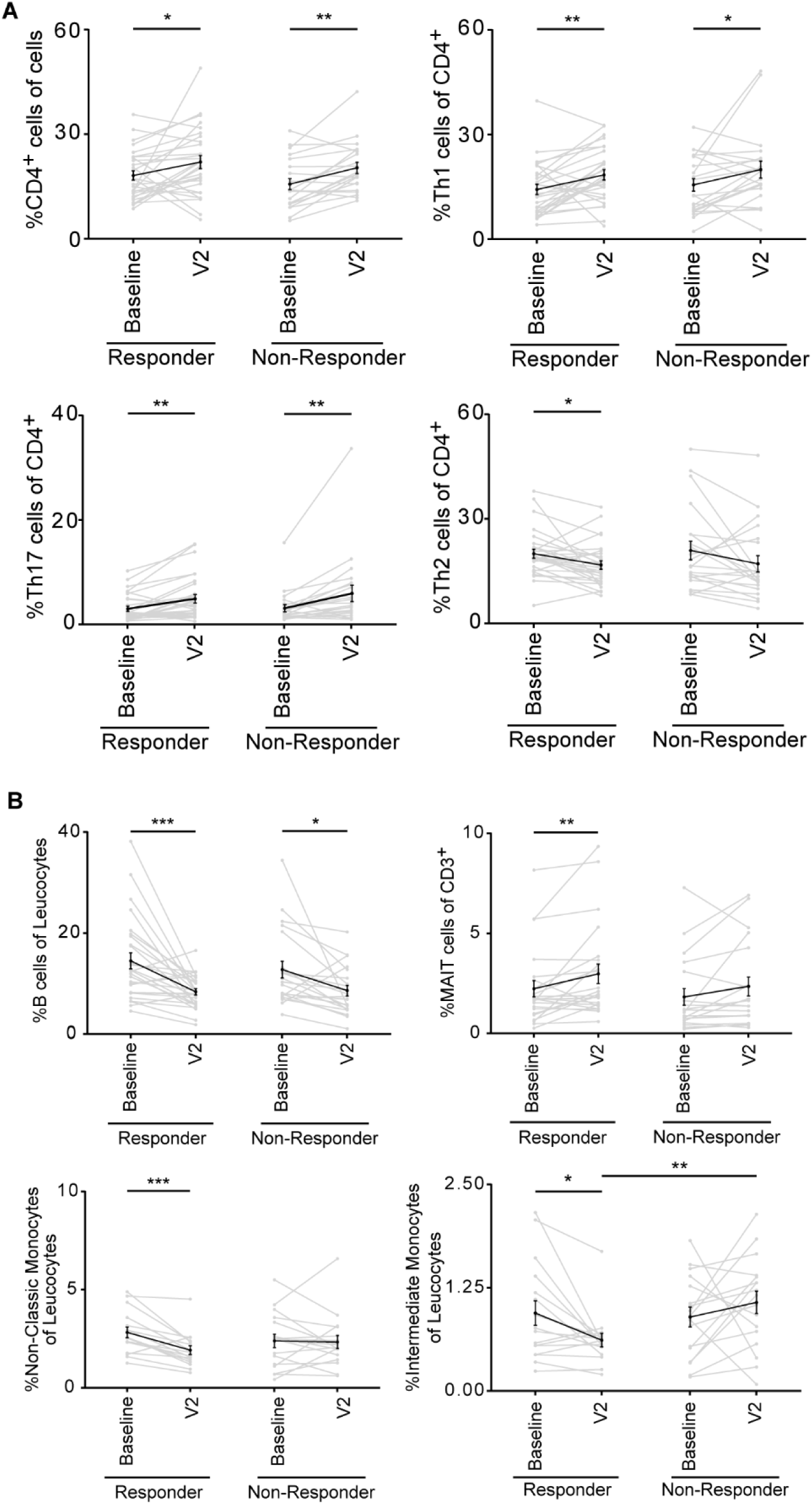
Supplementation differentially affects immune cell subsets in responding patients. (**A**) Proportions of CD4-, Th1-, Th17- and Th2 T cells among PBMCs, depicted as median split MDS-UPDRS III responder/nonresponder, matched baseline and V2, mean ± SEM. Significance was determined by paired T test. (**B**) Proportions of B cells, MAIT, nonclassic and intermediate monocyte populations in PBMCs depicted as median split MDS-UPDRS III responder/nonresponder, matched baseline and V2, mean ± SEM. Significance was determined by paired T test. *** p < 0.001, ** p < 0.01, *p < 0.05.

### Multiobjective analysis reveals parameters associated with best response to intervention

We initially defined responders and nonresponders by a reduction in MDS-UPDRS III after 6 months of supplementation. However, this method did not consider patients showing improvement in olfactory and/or cognitive function or improvement in one score but worsening in another score. Therefore, we devised an MOA to incorporate the various clinical measurements and thus more accurately determine the response to intervention. We performed nondominated sorting on the clinical parameter difference (V2 – baseline) with three metrics (objectives) olfactory score, MDS-UPDRS III and PANDA. We selected the 20% of patients who had the lowest and highest front numbers and sorted them into two clusters. These clusters included patients with the best/worst response to intervention (Fig. 4A). One major feature in our dataset concerns the different scales and distributions of the parameters, which makes it difficult to use existing statistical testing methodologies^24^. Therefore, we performed binary correlation-based feature selection to identify the physiological parameters that have the strongest correlation with the best/worst response to intervention. Using these two values, we identified the parameters with high levels of deviation that also had low levels of variation within a cluster (Fig. 4B). This approach enabled the identification of parameters associated with the best or worst combined clinical response (Fig. 4C). This novel approach of MOA revealed additional immune cell subsets associated with response to intervention. We identified CD4 Tfh cells as the parameter with the highest correlation to clinical response (Fig. 4D). This association was not evident when immune cell data were stratified by median split on MDS-UPDRS III (Fig. 4E). Thus, MOA is a promising novel approach to identify parameters associated with response to intervention in complex datasets with several conflicting metrics describing clinical outcome and a highly diverse repertoire of metrics recorded as potential correlates.

**Fig. 4.**
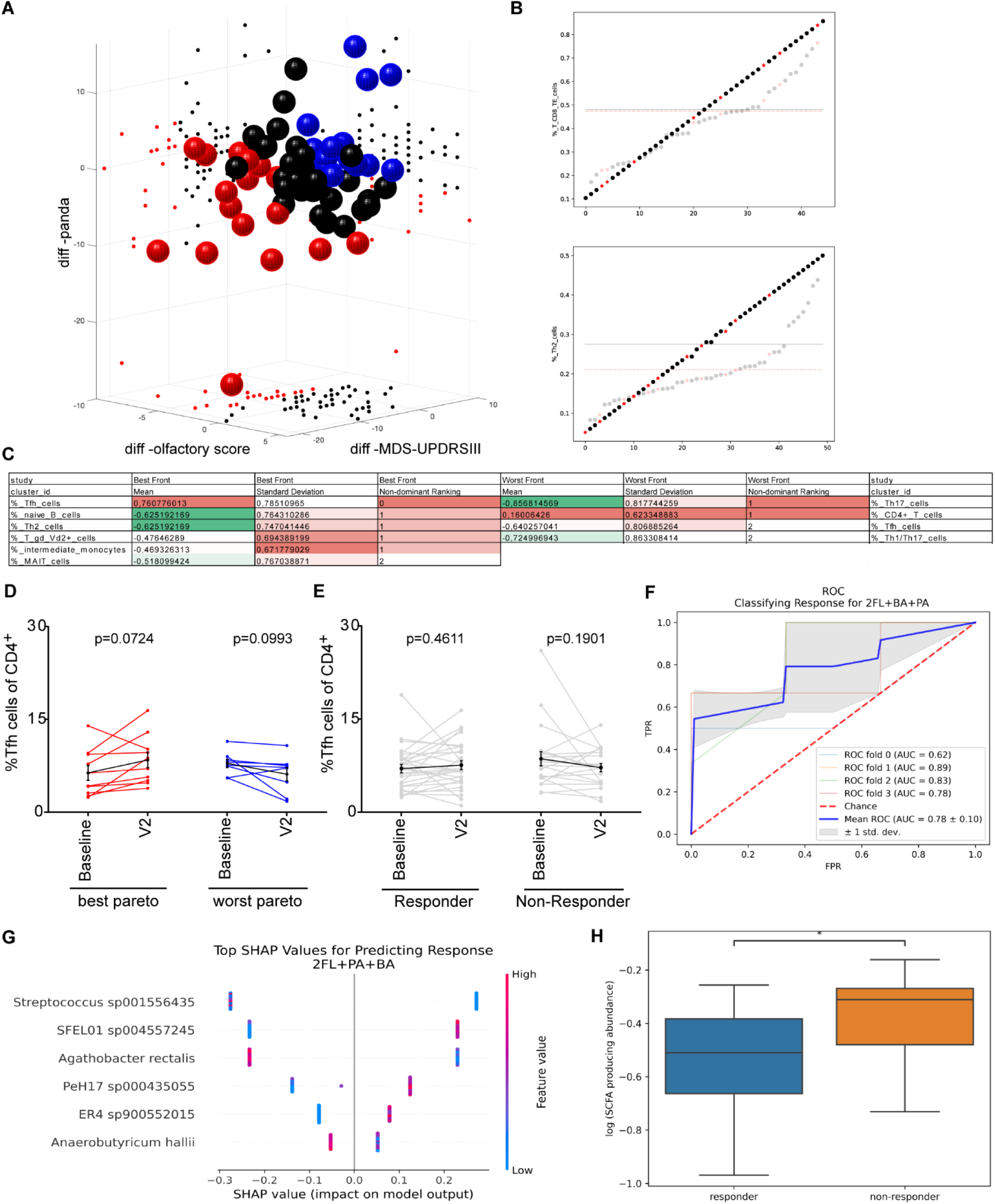
Multiobjective analysis and prediction modeling reveal parameters associated with the best/worst response to intervention. (**A**) Nondominated sorting based on olfactory score, MDS-UPDRS III and PANDA (V2 – baseline). The clusters of patients with the best (red) and worst (blue) 20% performances. (**B**) Examples of an uncorrelated (upper panel) and a correlated parameter (lower panel). Solid points depict ranking (not to scale), and transparent points the raw values, data points in the best cluster (red). Mean of all rankings (solid black line) and mean only in the best cluster (dashed red line). (**C**) Nondominated ranking of physiological parameters associated with the best/worst response. (**D**) Proportions of CD4 Tfh cells from patients within the best and worst clusters, matched baseline and V2, black line mean ± SEM. (**E**) Proportions of CD4 Tfh cells in all patients, median split MDS-UPDRS III responder/nonresponder, matched baseline and V2, black line mean ± SEM. Significance was determined by paired T test. (**F**) Prediction model for distinguishing R from NR: ROC curve of a prediction model based solely on microbiome baseline features (blue). (**G**) SHAP analysis of the model. (**H**) Abundances of SCFA-producing bacteria before supplementation with 2FL+BA+PA, *p < 0.05.

### PD patients’ microbiome before 2FL+BA+PA supplementation is associated with response

To assess whether the response to supplementation was affected by the patients’ microbiome, we used baseline microbiome abundances, as they may hold a potential to predict the impact of intervention. We devised a prediction model utilizing an XGBoost classifier. The model solely uses baseline microbiome data as its inputs and outputs a prediction of the treatment response. The model based on the microbiome of patients who received only 2FL or BA+PA had no predictive capability. However, the model predicted response when it was established based on the baseline microbiome of participants who received 2FL+BA+PA (Fig. 4F). We conducted SHapley Additive exPlanations (SHAP) analysis (Fig. 4G) to understand which species had the greatest impact on the model’s prediction. Our analysis revealed that *Streptococcus* sp001556435 and *Agathobacter rectalis* contributed to the prediction of nonresponders, whereas SFEL01 sp004557245 had a significant impact on the prediction of responders. *Agathobacter rectalis* is a SCFA-producing bacterium. Our study also revealed that the total abundance of SCFA-producing bacteria was significantly different between responders and nonresponders who were treated with 2FL+BA+PA; responders had a lower abundance of SCFA-producing bacteria (Fig. 4H).

## DISCUSSION

Here, we report that supplementation with SCFA improves clinical outcome in PD patients in a randomized double-blind 6-month prospective study with 72 patients. To our knowledge, this is the first exploratory study of SCFAs in PD patients showing improvement in motor and nonmotor functions beyond the level provided by standard medication.

Collectively, improvements in hyposmia, motor and cognitive function as a result of SCFA supplementation show that the intervention has a systemic impact on multiple brain regions. Amelioration of motor and nonmotor symptoms suggests a neuroregenerative element of SCFA supplementation. While we were not able to collect cerebrospinal fluid (CSF) in this study, we have previously shown increased PA concentrations in CSF after supplementation in MS patients.^10^ In addition, free fatty acid receptor 3 is expressed on the human brain endothelium, and its interaction protects the BBB from oxidative stress.^25^ Thus, supplementation with SCFAs may also directly impact the BBB and CNS to promote neuroregeneration.

Supplementation did not alter microbial composition. Even though microbial dysbiosis has been reported in PD patients, our results suggest that amelioration of disease symptoms by SCFA and/or 2FL supplementation does not require profound changes in gut microbe populations. The SCFA concentration in fecal samples also remained unaffected by supplementation, suggesting that excess SCFA is not excreted via stool. Over 95% of SCFAs produced in the colon are absorbed by the gut mucosa in healthy individuals^26^ or transported via the portal vein to the liver, preventing escape into the systemic circulation.^27^ Since SCFAs are not passed in the stool and serum levels only marginally increase, SCFAs are most likely swiftly metabolized.

Supplementation led to a significant increase in MAIT cells only in responding patients. At barrier surfaces, MAIT cells play an important role in the crosstalk between the host and the microbiome.^28^ Importantly, a regulatory role for MAIT cells in neuroinflammation through suppression of pathogenic Th1 cells has been shown,^29^ and reduced numbers of MAIT cells in peripheral blood have been reported in patients with several autoimmune diseases.^30^ A shift toward proinflammatory states in monocyte populations with a decrease in the levels of classic subsets and an increase in the levels of intermediate subsets with increased HLA-DR expression has been shown in PD,^31^ and a large study on expression quantitative trait loci (eQTL) revealed the overrepresentation of PD-related genes in monocyte populations.^32^ Decreased absolute counts of Tfh cells have also recently been reported in PD patients,^33^ in addition to decreased numbers of circulating anti-inflammatory B-cell subsets, thus implicating defective B-cell regulation as another contributing factor in PD. In summary, we propose that supplementation with SCFAs acts in concert on innate-like lymphoid cells, monocytes and B cells to promote a reduced inflammatory immune response in PD patients.

Our prediction model suggests that the patient microbiome can predict the efficacy of 2FL and BA+PA supplementation. Specifically, the composition of the SCFA-producing gut microbiota affected the response, with responders having less SCFA-producing gut microbes, which could help explain why supplementation with SCFA induces a greater response rate among them. If validated in larger studies, such a model can help evaluate the chances of a specific patient benefitting from intervention before it is started. In summary, SCFA supplementation may be a promising disease-modifying strategy in PD, hence, a follow-up phase III clinical trial to investigate the therapeutic potential of SCFAs in PD is warranted.

## Data Availability

All new datasets will be made freely available at the time of publication. Any additional information required to reanalyze the data reported in this work is available from the corresponding author upon request. This paper does not report any original code or algorithms. The code used in this study is freely available at https://www.ci.ovgu.de/Research/Codes.html and on GitHub.

## Acknowledgment

We thank Cathrine Husberg from BASF for her scientific contribution and energetic support. We thank Bärbel Henke from the Clinic of Neurology II, Hattingen for archiving the biomaterials. We thank Grazyna Debska-Vielhaber from the Department of Neurology, Magdeburg, for performing the seahorse assay. The help of Roland Hartig from the multiparametric bioimaging and cytometry platform (MPBIC) of the Otto-von-Guericke University Magdeburg with FACSAria cell sorting is greatly acknowledged.

## Author’s Roles

Conceptualization: TH, AD, CD, UOJ, RG, HP, NY, AH

Methodology: TH, AD, CD, GIS, FH, SK, AM, NY, AH

Software: MS, QS, SM, ES, NY

Formal Analysis: TH, AD, CD, SF, MS, QS, GIS, FH, SK, AM, CAD, NT, AZ, IES, SM, ES, NY, AH

Investigation: TH, AD, CD, SF, MS, FH, AM, LMW, JP, DJ, CAD

Resources: UOJ, SOH

Data Curation: TH, AD, CD, SF, MS, QS, LMW, JP, DJ, SM, ES, NY, AH

Writing – Original Draft: TH, AD, CD, NY, AH

Writing – Review & Editing: TH, AD, CD, MS, GIS, AZ, IES, SM, HP, ES, NY, AH

Visualization: TH, AD, CD, SF, MS, QS, NY, AH

Supervision: RG, HP, ES, NY, AH

Project Administration: TH, AD, CD, HP, AH

Funding Acquisition: UOJ, SOH, HP, AH

---

**Supplementary Fig. S1.**
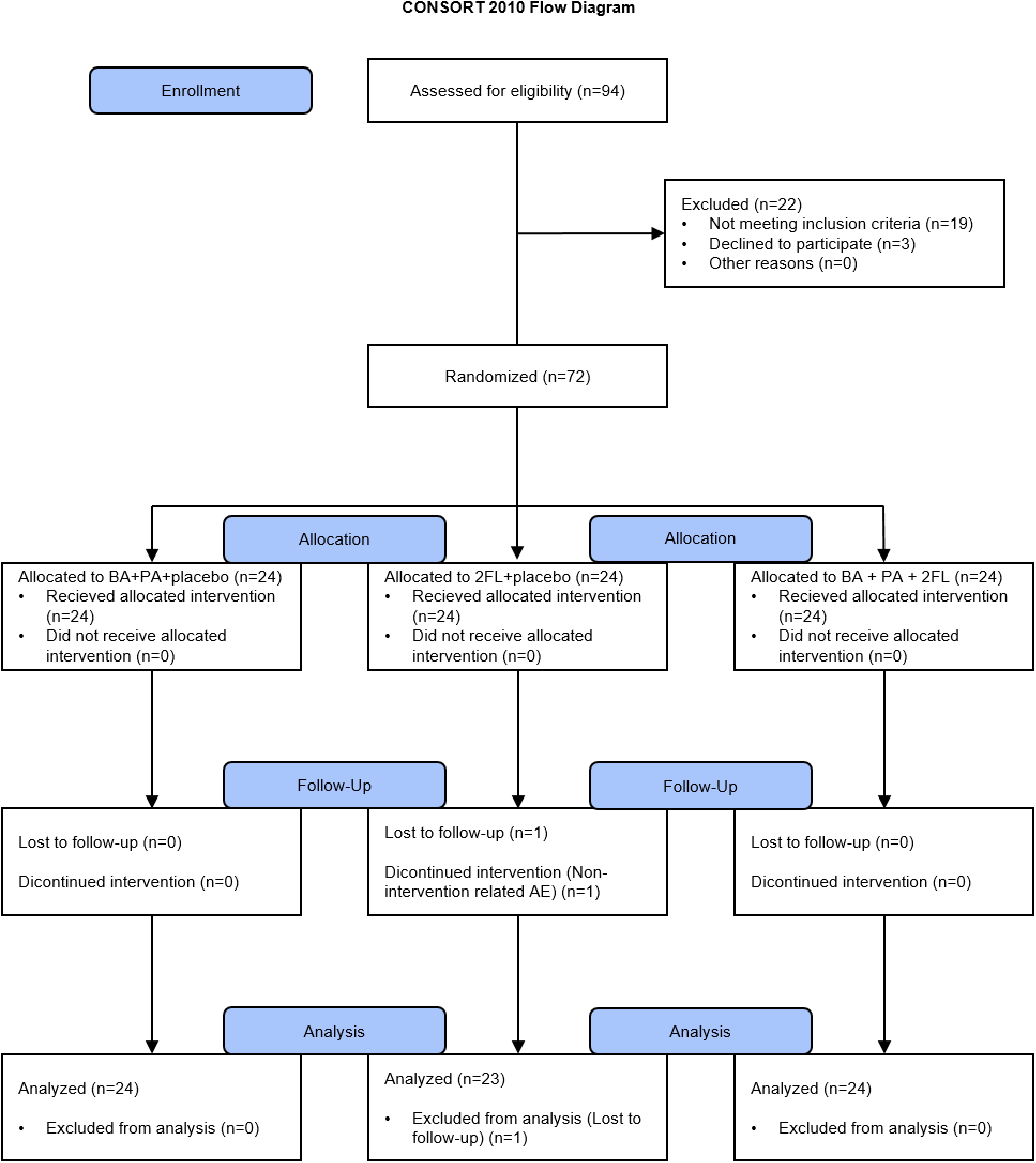
CONSORT 2010 flow diagram. CONsolidated Standards of Reporting Trials: Schematic representation of the randomized double-blind prospective study.

**Supplementary Fig. S2.**
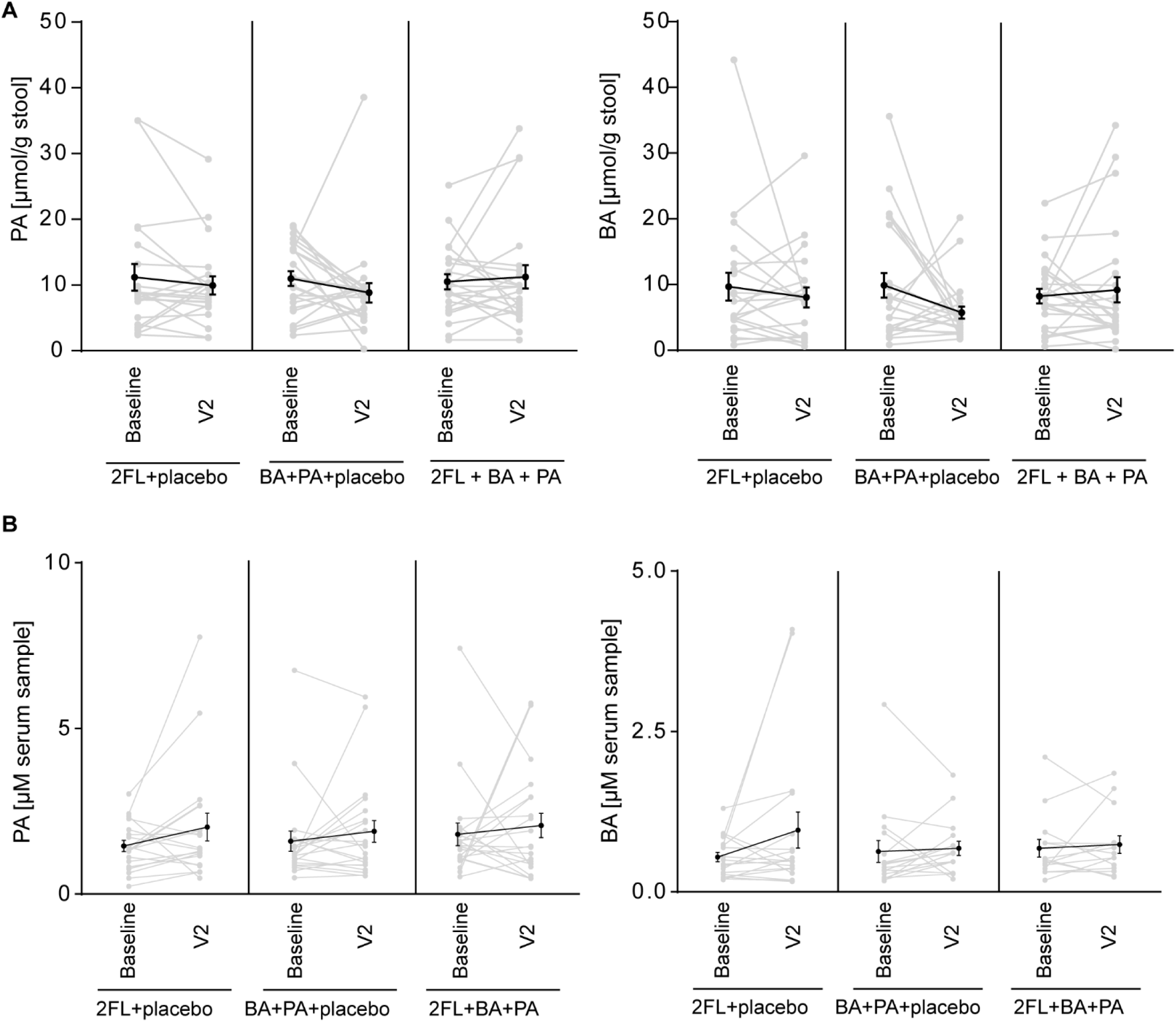
SCFA concentration in stool and serum is not altered after supplementation. Concentrations of PA and BA in stool (**A**) and in serum (**B**) samples measured by HPLC‒MS/MS. Data are plotted as before-after for each patient (gray) and mean (black) +/- SEM. Tested by 2-way ANOVA with Sidak’s multiple comparisons test.

**Supplementary Fig. S3:**
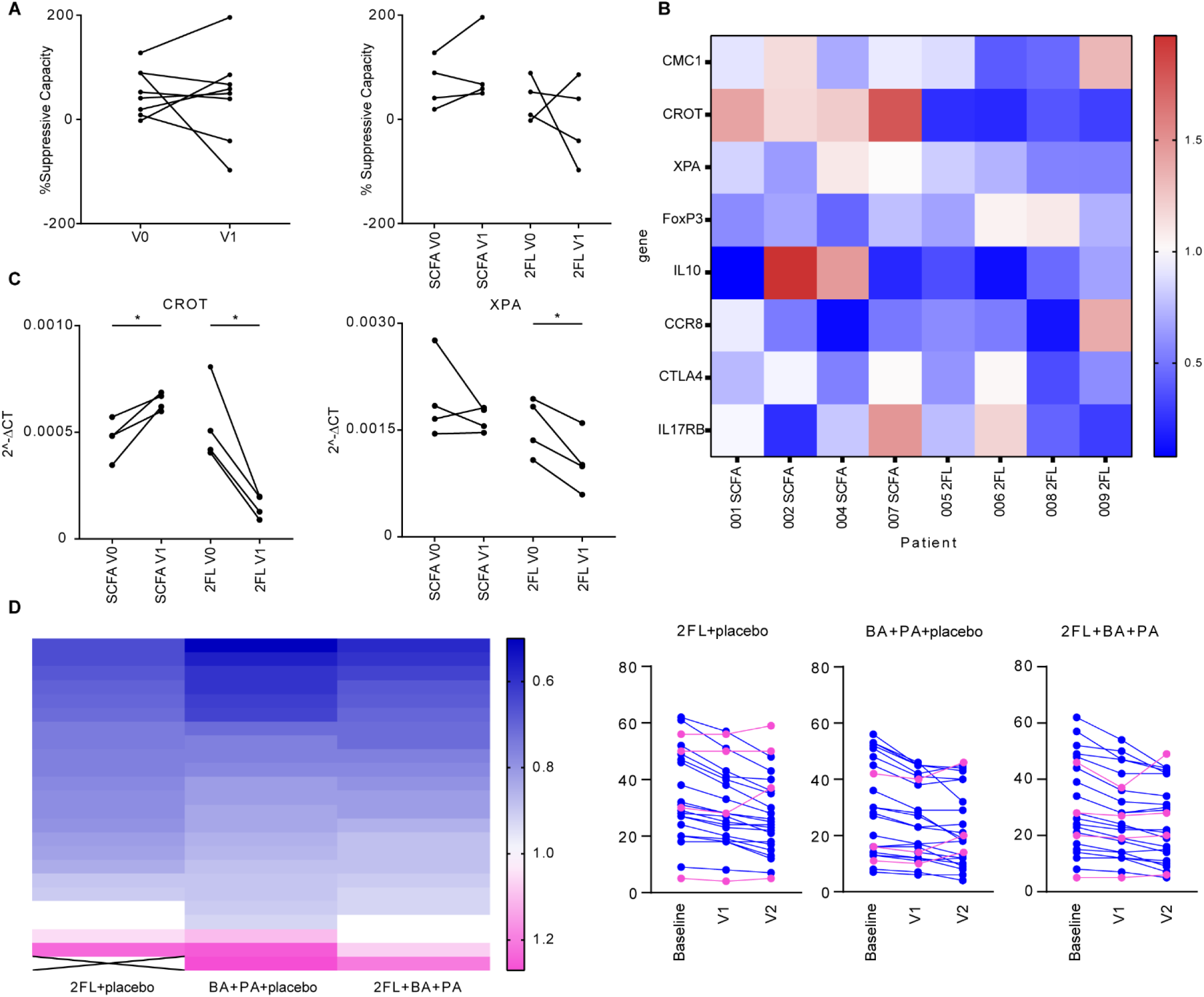
Supplementation with SCFA does not boost the suppressive capacity of Tregs but increases the expression of mitochondrial genes. (**A**) Treg/PBMC suppression assay at baseline (V0) and 14-day follow-up (V1), samples from validation cohort; MACS-sorted cells. (**B, C**) Gene expression in sorted Tregs from validation cohort; FACS-sorted cells. (**B**) Heatmap of fold changes follow-up (V1) and (**C**) individual 2^-ΔCT values for Crot and Xpa. Housekeeping B2m, time points compared by Wilcoxon matched pairs test. (**D**) Responders (R: MDS-UPDRS III V2 < MDS-UPDRS III baseline) and nonresponders (NR: MDS-UPDRS III V2 ≥ MDS-UPDRS III baseline) stratified by group. Data plotted as heatmap (ratio V2/baseline) and MDS-UPDRS III before-after for each patient; responders (blue) and nonresponders (pink).

**Supplementary Table S1:** List of antibodies used for in-depth immunophenotyping.

| Surface Marker | Clone | Fluorochrome | Company | Cat-Nr. | Panel |
| --- | --- | --- | --- | --- | --- |
| CD3 | UCHT1 | FITC | BioLegend | 300440 | 1, 2, 4 |
| CD4 | RPAT4 | APC Fire 750 | BioLegend | 300560 | 1 |
| CD25 | M-A251 | BV785 | BioLegend | 356140 | 1 |
| CD127 | A019D5 | APC | BioLegend | 351316 | 1 |
| CXCR5 | J25204 | PE/Dazzle 594 | BioLegend | 356928 | 1 |
| CD45RA | H100 | BV605 | BioLegend | 304134 | 1,2 |
| CCR7 | G043H7 | PerCP/Cy5.5 | BioLegend | 353220 | 1,2 |
| CCR6 | 11A9 | PE | BD | 559562 | 1 |
| CXCR3 | G025H7 | BV650 | BioLegend | 353730 | 1 |
| CCR4 | L2A1H4 | BV421 | BioLegend | 359414 | 1 |
| CD8 | SK1 | APC Fire 750 | BioLegend | 344746 | 2 |
| CD161 | HP3G10 | PE | BioLegend | 339904 | 2 |
| Vα7.2 | 3C10 | BV785 | BioLegend | 351722 | 2 |
| TCR $\gamma/\delta$ | 11F2 | APC | Miltenyi | 130-113-500 | 2 |
| Vα2 | B6 | BV421 | BioLegend | 331428 | 2 |
| CD19 | H1B19 | FITC | BioLegend | 302206 | 3, 4 |
| IgD | 1A6-2 | PE/Dazzle 594 | BioLegend | 348240 | 3 |
| CD45 | H130 | Alexa Fluor 700 | BioLegend | 304024 | 3, 4 |
| CD27 | 323 | BV421 | BioLegend | 302824 | 3 |
| CD38 | HIT2 | APC | BioLegend | 303510 | 3 |
| CD34 | 563 | PE | BD | 550761 | 3 |
| CD11c | B-LY6 | APC | BD | 559877 | 4 |
| CD14 | HCD14 | PE/Dazzle 594 | BioLegend | 325634 | 4 |
| CD16 | 3G8 | APC Fire 750 | BioLegend | 302060 | 4 |
| CD56 | HCD56 | PerCP/Cy5.5 | BioLegend | 318322 | 4 |
| CD11b | ICRF44 | PE | BioLegend | 301306 | 4 |
| CD123 | 6H6 | BV605 | BioLegend | 306026 | 4 |
| HLA-DR | L243 | BV785 | BioLegend | 307642 | 4 |

**Supplementary Table S2:**
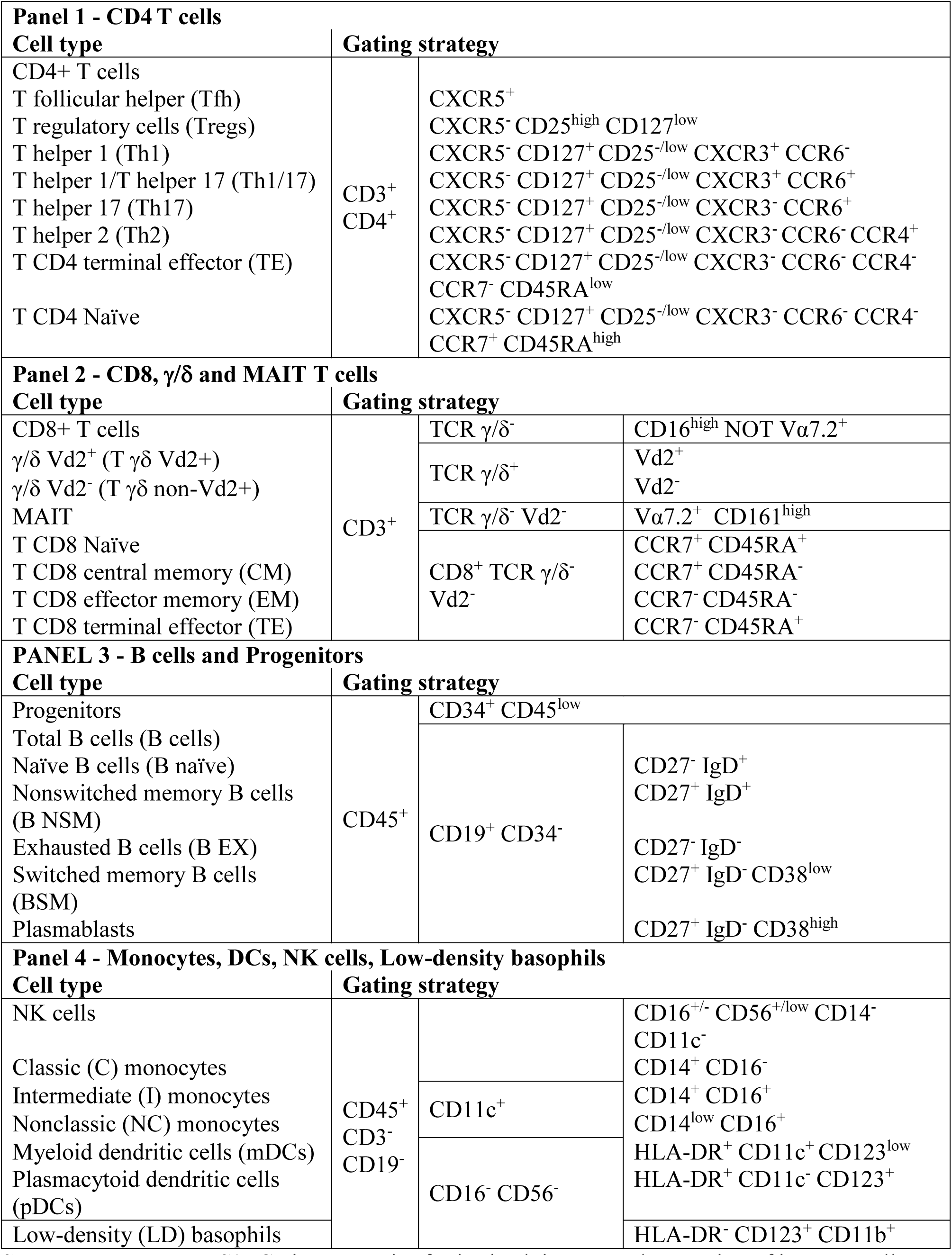
Gating strategies for in-depth immuno-phenotyping of immune cell subsets. Adapted from Monaco et al.^17^.

**Supplementary Table S3.** Demographic data and characteristics of patients prior to therapeutic intervention. Main demographics and clinical and treatment characteristics of participants at study entry. Data are presented as the mean ± SD or n (%).

| Characteristics | PA+BA+placebo<br>(n=24) | 2FL+placebo<br>(n=24) | 2FL+PA+BA<br>(n=24) |
| --- | --- | --- | --- |
| Female sex | 13 (54.2%) | 9 (37.5%) | 13 (54.2%) |
| Age, years | 62(±9) | 67 (±12) | 65 (±8) |
| Duration of disease, years | 7 (±7) | 6 (±6) | 4 (±4) |
| BMI | 26 (±4) | 26 (±3) | 25 (±3) |
| Subgroups |  |  |  |
| akinetic-rigid | 8 (33.3%) | 9 (37.5%) | 11 (45.8%) |
| equivalent | 14 (58.3%) | 12 (50%) | 12 (50%) |
| tremordominant | 2 (8.3 %) | 3 (12.5%) | 1 (4.17%) |
| Medication |  |  |  |
| LEDD [mg] | 770 (±302) | 785 (±267) | 756 (±261) |
| 95% CI | 642.1-897.1 | 672.2-897.3 | 646-866.5 |
| Benserazide | 24 (100%) | 22 (91.7%) | 23 (95.8%) |
| Carbidopa | 4 (16.7%) | 5 (20.8%) | 2 (8.3%) |
| Piribedil | 2 (8.3%) | 0 | 2 (8.3%) |
| Rotigotine | 2 (8.3%) | 3 (12.5%) | 3 (12.5%) |
| Ropinirole | 2 (8.3%) | 1 (4.17%) | 2 (8.3%) |
| Pramipexole | 4 (16.7%) | 2 (8.3%) | 4 (16.7%) |
| Rasagiline | 13 (54.2%) | 8 (33.3%) | 14 (58.3%) |
| Selegiline | 3 (12.5%) | 9 (37.5%) | 3 (12.5%) |
| Safinamide | 2 (8.3%) | 1 (4.2%) | 2 (8.3%) |
| COMT-inhibitor | 6 (25%) | 8 (33.3%) | 9 (37.5%) |
| Amantadine | 2 (8.3 %) | 2 (8.3 %) | 1 (4.2%) |
| Apomorphine | 0 | 0 | 0 |
| Anticholinergics | 0 | 1 (4.2%) | 0 |
| Clozapine | 1 (4.2%) | 0 | 2 (8.3%) |
| Quetiapine | 1 (4.2%) | 3 (12.5%) | 1 (4.2%) |
| MDS-Unified Parkinson's Disease<br>Rating Scale |  |  |  |
| MDS-UPDRS I | 10 (±10) | 17 (±12) | 17 (±13) |
| 95% CI | 12.6-21 | 12.1-22.5 | 11.2-22.1 |
| MDS-UPDRS II | 9 (±6) | 9 (±8) | 10 (±9) |
| 95% CI | 6.8-12.1 | 6.3-12.7 | 6-13.4 |
| MDS-UPDRS III | 29 (±17) | 33 (±16) | 30 (±16) |
| 95% CI | 22.1-36.3 | 26.3-40 | 23-36.8 |
| MDS-UPDRS IV | 3 (± 3) | 5 (±5) | 3 (±3) |
| 95% CI | 1.8-4.2 | 2.5-6.3 | 1.4-4.1 |
| Hoehn & Yahr scale | 2 (±1) | 2 (±1) | 2 (±1) |
| 95% CI | 1.9-2.5 | 1.9-2.4 | 1.9-2.7 |
| PANDA | 25 (±4) | 33 (±16) | 30 (±16) |
| 95% CI | 23.7-26.7 | 18.9-24.8 | 20.8-26.1 |
| Olfactory score | 8 (±3) | 7 (±3) | 7 (±3) |
| 95% CI | 6.8-9 | 5.9-8.1 | 5.6-8.1 |

## REFERENCES

1. Poewe W, Seppi K, Tanner CM, et al. Parkinson disease. Nat Rev Dis Primers 2017;3:17013.

2. Schapira AHV, Chaudhuri KR, Jenner P. Non-motor features of Parkinson disease. Nat Rev Neurosci 2017;18(8):509.

3. Zinocker MK, Lindseth IA. The Western Diet-Microbiome-Host Interaction and Its Role in Metabolic Disease. Nutrients 2018;10(3).

4. Cryan JF, O’Riordan KJ, Sandhu K, Peterson V, Dinan TG. The gut microbiome in neurological disorders. Lancet Neurol 2020;19(2):179–194.

5. Forsyth CB, Shannon KM, Kordower JH, et al. Increased intestinal permeability correlates with sigmoid mucosa alpha-synuclein staining and endotoxin exposure markers in early Parkinson’s disease. PLoS One 2011;6(12):e28032.

6. Nie S, Wang J, Deng Y, Ye Z, Ge Y. Inflammatory microbes and genes as potential biomarkers of Parkinson’s disease. NPJ Biofilms Microbiomes 2022;8(1):101.

7. Romano S, Savva GM, Bedarf JR, Charles IG, Hildebrand F, Narbad A. Meta-analysis of the Parkinson’s disease gut microbiome suggests alterations linked to intestinal inflammation. NPJ Parkinsons Dis 2021;7(1):27.

8. Visconti A, Le Roy CI, Rosa F, et al. Interplay between the human gut microbiome and host metabolism. Nat Commun 2019;10(1):4505.

9. Unger MM, Spiegel J, Dillmann KU, et al. Short chain fatty acids and gut microbiota differ between patients with Parkinson’s disease and age-matched controls. Parkinsonism Relat Disord 2016;32:66–72.

10. Duscha A, Gisevius B, Hirschberg S, et al. Propionic Acid Shapes the Multiple Sclerosis Disease Course by an Immunomodulatory Mechanism. Cell 2020;180(6):1067–1080 e1016.

11. Hall DA, Voigt RM, Cantu-Jungles TM, et al. An open label, non-randomized study assessing a prebiotic fiber intervention in a small cohort of Parkinson’s disease participants. Nat Commun 2023;14(1):926.

12. Suligoj T, Vigsnaes LK, Abbeele PVD, et al. Effects of Human Milk Oligosaccharides on the Adult Gut Microbiota and Barrier Function. Nutrients 2020;12(9).

13. Leviatan S, Shoer S, Rothschild D, Gorodetski M, Segal E. An expanded reference map of the human gut microbiome reveals hundreds of previously unknown species. Nat Commun 2022;13(1):3863.

14. Chen T, Guestrin C. XGBoost: A Scalable Tree Boosting System. Proceedings of the 22nd ACM SIGKDD International Conference on Knowledge Discovery and Data Mining. San Francisco, California, USA: Association for Computing Machinery; 2016. p. 785–794.

15. Wang XW, Liu YY. Comparative study of classifiers for human microbiome data. Med Microecol 2020;4.

16. Lundberg SM, Erion GG, Lee S-I. Consistent Individualized Feature Attribution for Tree Ensembles. ArXiv 2018;abs/1802.03888.

17. Monaco G, Lee B, Xu W, et al. RNA-Seq Signatures Normalized by mRNA Abundance Allow Absolute Deconvolution of Human Immune Cell Types. Cell Rep 2019;26(6):1627–1640 e1627.

18. Deb K, Agrawal S, Pratap A, Meyarivan T. A Fast Elitist Non-dominated Sorting Genetic Algorithm for Multi-objective Optimization: NSGA-II. 2000; Berlin, Heidelberg: Springer Berlin Heidelberg. p. 849–858.

19. Goetz CG, Tilley BC, Shaftman SR, et al. Movement Disorder Society-sponsored revision of the Unified Parkinson’s Disease Rating Scale (MDS-UPDRS): scale presentation and clinimetric testing results. Mov Disord 2008;23(15):2129–2170.

20. Tomlinson CL, Stowe R, Patel S, Rick C, Gray R, Clarke CE. Systematic review of levodopa dose equivalency reporting in Parkinson’s disease. Mov Disord 2010;25(15):2649–2653.

21. Cavaco S, Goncalves A, Mendes A, et al. Abnormal Olfaction in Parkinson’s Disease Is Related to Faster Disease Progression. Behav Neurol 2015;2015:976589.

22. Michels J, van der Wurp H, Kalbe E, et al. Long-Term Cognitive Decline Related to the Motor Phenotype in Parkinson’s Disease. J Parkinsons Dis 2022;12(3):905–916.

23. Correa-Oliveira R, Fachi JL, Vieira A, Sato FT, Vinolo MA. Regulation of immune cell function by short-chain fatty acids. Clin Transl Immunology 2016;5(4):e73.

24. Lualdi M, Fasano M. Statistical analysis of proteomics data: A review on feature selection. J Proteomics 2019;198:18–26.

25. Hoyles L, Snelling T, Umlai UK, et al. Microbiome-host systems interactions: protective effects of propionate upon the blood-brain barrier. Microbiome 2018;6(1):55.

26. Salminen S, Bouley C, Boutron-Ruault MC, et al. Functional food science and gastrointestinal physiology and function. Br J Nutr 1998;80 Suppl 1:S147–171.

27. Bloemen JG, Venema K, van de Poll MC, Olde Damink SW, Buurman WA, Dejong CH. Short chain fatty acids exchange across the gut and liver in humans measured at surgery. Clin Nutr 2009;28(6):657–661.

28. Amini A, Pang D, Hackstein CP, Klenerman P. MAIT Cells in Barrier Tissues: Lessons from Immediate Neighbors. Front Immunol 2020;11:584521.

29. Miyazaki Y, Miyake S, Chiba A, Lantz O, Yamamura T. Mucosal-associated invariant T cells regulate Th1 response in multiple sclerosis. Int Immunol 2011;23(9):529–535.

30. Mechelli R, Romano S, Romano C, et al. MAIT Cells and Microbiota in Multiple Sclerosis and Other Autoimmune Diseases. Microorganisms 2021;9(6).

31. Thome AD, Atassi F, Wang J, et al. Ex vivo expansion of dysfunctional regulatory T lymphocytes restores suppressive function in Parkinson’s disease. NPJ Parkinsons Dis 2021;7(1):41.

32. Raj T, Rothamel K, Mostafavi S, et al. Polarization of the effects of autoimmune and neurodegenerative risk alleles in leukocytes. Science 2014;344(6183):519–523.

33. Li R, Tropea TF, Baratta LR, et al. Abnormal B-Cell and Tfh-Cell Profiles in Patients With Parkinson Disease: A Cross-sectional Study. Neurol Neuroimmunol Neuroinflamm 2022;9(2).

